# Potential involvement of monoamine oxidase activity in delirium onset and SARS-COV2 infection

**DOI:** 10.1101/2020.06.16.20128660

**Authors:** Miroslava Cuperlovic-Culf, Emma L. Cunningham, Anu Surendra, Xiaobei Pan, Steffany A.L. Bennett, Mijin Jung, Bernadette McGuiness, Anthony Peter Passmore, Danny McAuley, David Beverland, Brian D. Green

## Abstract

Delirium is an acute change in attention and cognition occurring in ~65% of severe SARS-CoV-2 cases. It is also common following surgery and an indicator of brain vulnerability and risk for the development of dementia. In this work we analyzed the underlying role of metabolism in delirium-susceptibility in the postoperative setting using metabolomic profiling of cerebrospinal fluid and blood taken from the same patients prior to planned orthopaedic surgery. Significant concentration differences in several amino acids, acylcarnitines and polyamines were found in delirium-prone patients leading us to a hypothesis about the significance of monoamine oxidase B (MAOB) in predisposition to delirium. Subsequent computational structural comparison between MAOB and angiotensin converting enzyme 2 as well as protein-protein docking analysis showed possibly strong binding of SARS-CoV-2 spike protein to MAOB resulting in a hypothesis that SARS-CoV-2 influences MAOB activity possibly lead to many observed neurological and platelet-based complications of SARS-CoV-2 infection. This proposition is possibly of significance for diagnosis, treatment and prevention of vulnerabilities causing delirium, dementias and severe COVID-19 response.

## Introduction

COVID-19 is an ongoing major global health emergency caused by severe acute respiratory syndrome coronavirus SARS-CoV-2. Patients admitted to hospital with COVID-19 show a range of features including fever, anosmia, acute respiratory failure, kidney failure and gastrointestinal issues and the death rate of infected patients it is estimated at 2.2% (Rothan and Byrareddy, 2020). Some early studies of hospitalized patients indicated that 20-30% of COVID-19 patients develop some form of delirium or mental status change rising to 60-70% for those patients with severe illness of all age groups (Mao et al, 2020; Helms et al. 2020). The exact mechanisms are not understood although a number of possible causes have been proposed including direct viral invasion, cerebrovascular involvement, hypoxia, pyrexia, dehydration, hyperinflammation (cytokine storm), medications or metabolic derangements (Mao et al, 2020; Helms et al. 2020). The X-ray structure of SARS-CoV-2 spike protein binding to ACE2 (Lan et al. 2020; Shang et al. 2020) as well as recently demonstrated specific structural features of SARS-CoV-2 spike protein (Gussow et al. 2020), suggest specific features of SARS-CoV-2 spike protein structure for binding to human protein in the SARS-CoV-2 influence virulence.

Delirium, an acute disorder of attention and cognition (American Psychiatric Association, 2013) is an unpleasant experience for patients, relatives and healthcare staff and is associated with negative outcomes such as dementia and death. As has been described by Fong et al. (2015), whilst even those with the most resilient of brains can develop delirium in the face of severe stressors, delirium in the face of more moderate insults may be a sign of underlying neurodegeneration (Cunningham et al. 2019). An improved understanding of the causes of postoperative delirium could provide a better appreciation of the vulnerabilities causing delirium following surgery, as well as following SARS-CoV-2 infection.

Planned orthopedic surgery under spinal anaesthesia provides a unique opportunity to preoperatively sample cerebrospinal fluid (CSF) in a group of patients where an estimated 17% will subsequently develop delirium (Scott et al 2015). Metabolomic analysis of body fluids, including blood and CSF, provides a wide-ranging molecular window into the major processes of the body including an insight into the brain metabolism. CSF metabolite biomarkers of delirium risk have already been identified (Pan et al. 2019). Major differences in the concentrations of polyamines (including spermidine and putrescine) are present in delirium-prone patients even before surgery or the presentation of delirium. However, it is not clear whether such metabolic changes occur more peripherally in the blood circulation, or whether the transfer of these metabolite across the blood-brain barrier is important.

Recently, Shen et al. (Shen et al. 2020) presented metabolomics and proteomics analysis of serum of mild and severe COVID-19 patients indicating major concentration differences in serotonin, kynurenine, a number of amino acids, as well as, alterations in tryptophan and polyamine metabolic pathways. This metabolomics and proteomics analysis also indicated major role for platelets in SARS-CoV-2 infection. The metabolomics data were in agreement with clinical studies showing coagulopathy as one of the major issues in severe COVID-19 cases (Becker, 2020), Additionally, delirium, possibly associated with changes in neurotransmitters has been indicated as a severe consequence of SARS-CoV-2 infection in some patients (Alkeridy et al. 2020). The involvement of the enzyme monoamine oxidase (MAO) emerges as a common potential candidate causing many of the observed COVID-19 side-effects. MAO is important for neurotransmitter metabolism, has prior associations with delirium, and is involved in platelet regulation and coagulation (Deshwal et al. 2018; Leiter et al. 2020; Yeung et al. 2019) as well as anosmia (Ketzef et al. 2017). MAO including MAOA and MAOB are flavoenzymes that catalyze the oxidative transformation of monoamines. Inhibition of these enzymes is an established therapeutic target which is still in development, and the selectivity of candidate molecules for one isoform over another is a key consideration (Schedin-Weiss et al. 2017; Sharpe et al. 2016). Alterations in the activities of MAOs is a potential source of various neuropsychiatric disorders including depression, autism or aggressive behavior (Brunner et al. 1993; Shih et al. 1999). Additionally, function of MAOs represents an inherent source of oxidative stress, leading to the damage and death of neurons, ultimately leading to neurodegenerative diseases such as Parkinson’s or Alzheimer’s disease (Gandhi and Abramov, 2012; Naoi et al. 2011).

## Materials and Methods

### Samples and experimental analyses

Preoperative blood and CSF samples were collected within an observational cohort study of patients aged over 65 years without a diagnosis of dementia presenting for planned hip and knee replacements prior to the SARS-CoV-2 pandemic (approved by Office for Research Ethics Committee for Northern Ireland (REC ref: 10/NIR01/5)). The study methodology has been described in detail previously (Cunningham et al. 2019). The study population had a mean age of 74.4 years and 57% were female. The incidence of postoperative delirium was 14%. Paired CSF and blood metabolomics analysis was undertaken for 54 age and gender matched patients where half of the patients experienced post-operative delirium and half did not (control). Metabolomic analysis was undertaken where sufficient CSF was available for age and gender matched delirium and control participants.

### Metabolomics

The analysis of metabolic profiles of CSF samples from the nested case-control postoperative delirium cohort has previously been published (Pan et al. 2019). In this investigation the corresponding blood plasma samples of each of the same patients was examined by an identical kit-based methodology. Quantitative metabolomic profiling was performed using the Biocrates AbsoluteIDQ p180 (BIOCRATES, Life Science AG, Innsbruck, Austria) using a Xevo TQ-MS triple-quadrupole mass spectrometer (Waters Corporation, Milford, USA) as previously described (0 et al. 2016). Briefly, this comprised of two general methods: UPLC (I-Class, Waters Corporation, UK) reversed-phase (Waters ACQUITY UPLC BEH C18 2.1 × 50 mm, 1.7 μm; UK) with multiple reaction monitoring (MRM), and flow injection analysis (FIA) also operating in MRM mode. Metabolite concentrations were calculated and expressed as micromolar (μM).

### SARS-CoV-2 metabolomics dataset

Plasma metabolomics profiles of SARS-CoV-2 patients was published and made available by Shen et al. (Shen et al. 2020). The provided dataset includes metabolomic analysis of serum samples from patients with mild or severe COVID-19 as well as control subjects (with number of patient equal to mild 28, severe 37 and control 28). Briefly, authors used the ultra-performance liquid chromatography/tandem mass spectrometry (UPLC-MS/MS) for untargeted metabolomics of serum samples providing identification and quantification of 941 metabolites including 36 drugs and their metabolites. Details of methodology and validation are provided in the original publication (Shen et al. 2020).

### Data analysis and Protein simulations

Different machine learning and statistical methods running under Matlab 2020 (Matworks Inc), Orange 3.25 (Demsar et al. 2013), TMeV (Saeed et al. 2003) and Python – Jupyter Notebook were used for the analysis of metabolomics data. Principal component analysis (PCA) and 2-class Partial Least Squares-Discriminant Analysis (PLS-DA) model investigation was performed using jupyter notebook on data that was log10 transformed with selection to have an under 20% standard deviation and fewer than 10% of missing values in measurements for all samples. Selection of major features in different groups was performed using Significant Analysis of Microarrays (SAM) (Tusher and Tibshirani, 2001). Machine learning analysis was performed using Python and Matlab with the results of Python Random Forest classification with SHAP (SHapley Additive exPlanations) algorithm shown to explain the output of machine learning (Lundberg and Lee, 2017). A comparison of the protein structures of MAOB and ACE2 bonded to SARS-CoV-2 Spike protein are analyzed using UCSF Chimera, Schrodinger software package (Schrodinger Inc.) and PDB Data Bank structure analysis methodologies including jFATCAT (Ye and Godzik, 2003). Protein X-ray structures were obtained from https://www.rcsb.org/ and included for MAOB protein PDB ID 1GOS (Binda et al. 2001) and ACE2 bonded to SARS-CoV-2 Spike protein PDB ID: 6M0J.

Computational analysis of the spike-MAOB docking was performed using Schrodinger’s BioLuminate package with spike protein obtained from 6M0J PDB entry was used as ligand and each MAOB chain obtained from 6FWC PDB entry was used as a receptor. The opposite analysis with MAOB chains as ligands and Spike protein as receptor led to the same optimal result. MAOB membrane bound regions have been excluded in docking calculations.

## Results

### Detailed metabolomics analysis shows differences in Delirium-prone and Delirium-free patient groups prior to surgery in both blood and CSF metabolites

Principal component analysis (PCA) and 2-class Partial Least Squares-Discriminant Analysis (PLS-DA) were performed on CSF and blood metabolomic data samples. For both sample types PCA achieved only limited separation of control and delirium-prone cases (Supplementary Fig. 1A and 1B). Incorporation of patient gender had very minor improvement of PCA group separation (Supplementary Fig. 1E). Supervised PLS-DA identified combination of metabolite features capable of clearly distinguishing the two patient groups (Supplementary Fig. 1C and 1D). For the CSF data the output was closely aligned with the previously published analysis (Pan et al. 2019). The importance of specific metabolites to the model’s discriminatory power are shown in coefficient plots (Supplementary Fig. 2). The most important contributors in control and delirium sample separation in CSF were spermidine, putrescine, glutamine (as previously reported by Pan et al 2019), but also 3-hydroxypalmitoleylcarnitine (C16-1-OH), linoleic acid (C18-2), carnitine (C0) and, to a lesser extent, a number of other metabolites (Supplementary Fig. 2 shown in red). For blood metabolomic data the discrimination of control and delirium patients was influenced most by levels of proline, ornithine, lysine, trans-4-hydroxyproline (t4-OH-Pro), PC aa C24:0 and PC aa C26:0 as well as H1, ADMA, C7-DC and, to a lesser extent, a number of other metabolites (shown in red in Supplementary Fig. 2).

The above results were further corroborated by application of Statistical Analysis for Microarrays (SAM) (Tusher et al. 2001), which has been previously applied to metabolomics data (Cuperlovic-Culf et al. 2012) (Fig. 1A). SAM analysis of normalized CSF data found significant differences in CSF for ornithine, glutamine, putrescine, spermidine and threonine, similarly as previously shown (Pan et al. 2019) and in good agreement with the above PLS-DA (ornithine, glutamine, putrescine and spermidine were major features). Threonine was not one of the most significant contributors to the classification coefficients, but was still a significant contributor to the PLS-DA sample grouping. SAM analysis of blood metabolites shows major differences in pre-operative blood samples in proline, threonine, lysine, ornithine and phosphatidylcholines (specifically PC aa C26:0).

**Fig. 1.**
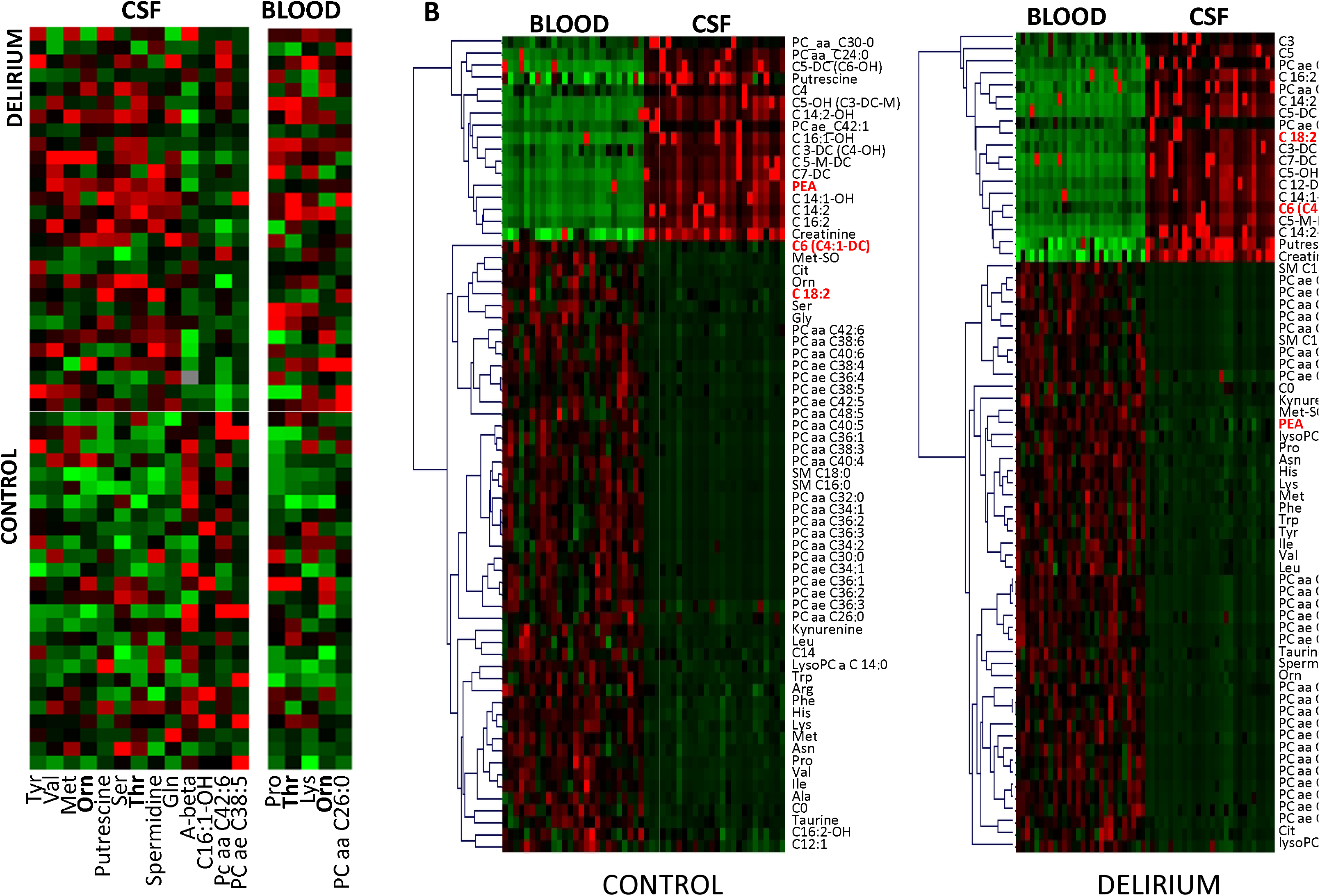
A. SAM selection of the most relevant metabolites for classification control and delirium group from metabolites in pre-operative CSF and blood samples; B SAM analysis of major differences between CSF and blood metabolites in the control and delirium groups. In both cases analysis was performed following metabolite and sample normalization in TMeV. For PEA – Control, adjusted p-value 1.8E-6; for PEA – Delirium, adjusted p-value 1.78 E-10.

### Phenethylamine, Octadecadienylcarnitine and hexanoylcarnitine show opposite concentration difference in blood and CSF in Delirium-prone and Delirium-free patient groups

The availability of metabolic profiles for CSF and blood provides a unique possibility for the determination of significant differences in metabolite concentrations between these two body fluids giving information about the transfer and metabolism across the blood brain barrier as well as the possibility for the determination of blood-based biomarkers that are representative of changes in the CSF (Fig. 1B). As can be expected there are major metabolic differences between CSF and blood in both control and delirium-prone group with majority of metabolites showing highly comparable behaviour in the two groups. Notable exceptions are metabolites that show opposite concentration in the two body fluids namely – phenethylamine (PEA) with higher concentration in CSF relative to blood in control subjects and opposite in delirium and Octadecadienylcarnitine (C 18:2) and hexanoylcarnitine (C6 (C4:1-DC)) showing higher concentration in blood then in CSF in control group. PEA is a natural monoamine alkaloid that acts as a central nervous system stimulant. Octadecadienylcarnitine (also called Linoleyl carnitine) and hexanoylcarnitine is a long-chain acyl fatty acid derivatives of carnitine. Difference in concentration in these three metabolites are significant in both control and delirium groups with adjusted p-values for blood to CSF groups in control: for PEA adj. p= 1.83e-6; C 18:2 adj. p= 6.1e-4 and C6 (C4:1-DC) adj. p= 6.6e-4 and delirium: PEA adj. p= 1.8e-10; C 18:2 adj. p= 2.8e-10 and C6 (C4:1-DC) adj. p= 5.9e-8. Although the difference in these three metabolites between CSF and blood is significant the difference of their concentrations between control and delirium observed separately in CSF and blood is only minor. In CSF PEA is overall slightly reduced in the delirium group (adj. p-value = 0.1) while C 18:2 and C6 (C4:1-DC) are unchanged. Similarly in plasma PEA concentration is slightly lower in delirium and C 18:2 and C6 (C4:1-DC) are unchanged. Therefore, overall PEA is slightly reduced in delirium-prone patients, prior to operation and significantly, in this group PEA concentration is significantly lower in CSF then in blood. PEA is a neurotransmitter integral to the signalling process in the central nervous system. It can be oxidised through the function of monoamine oxidase (MAO) enzymes.

**Fig. 2.**
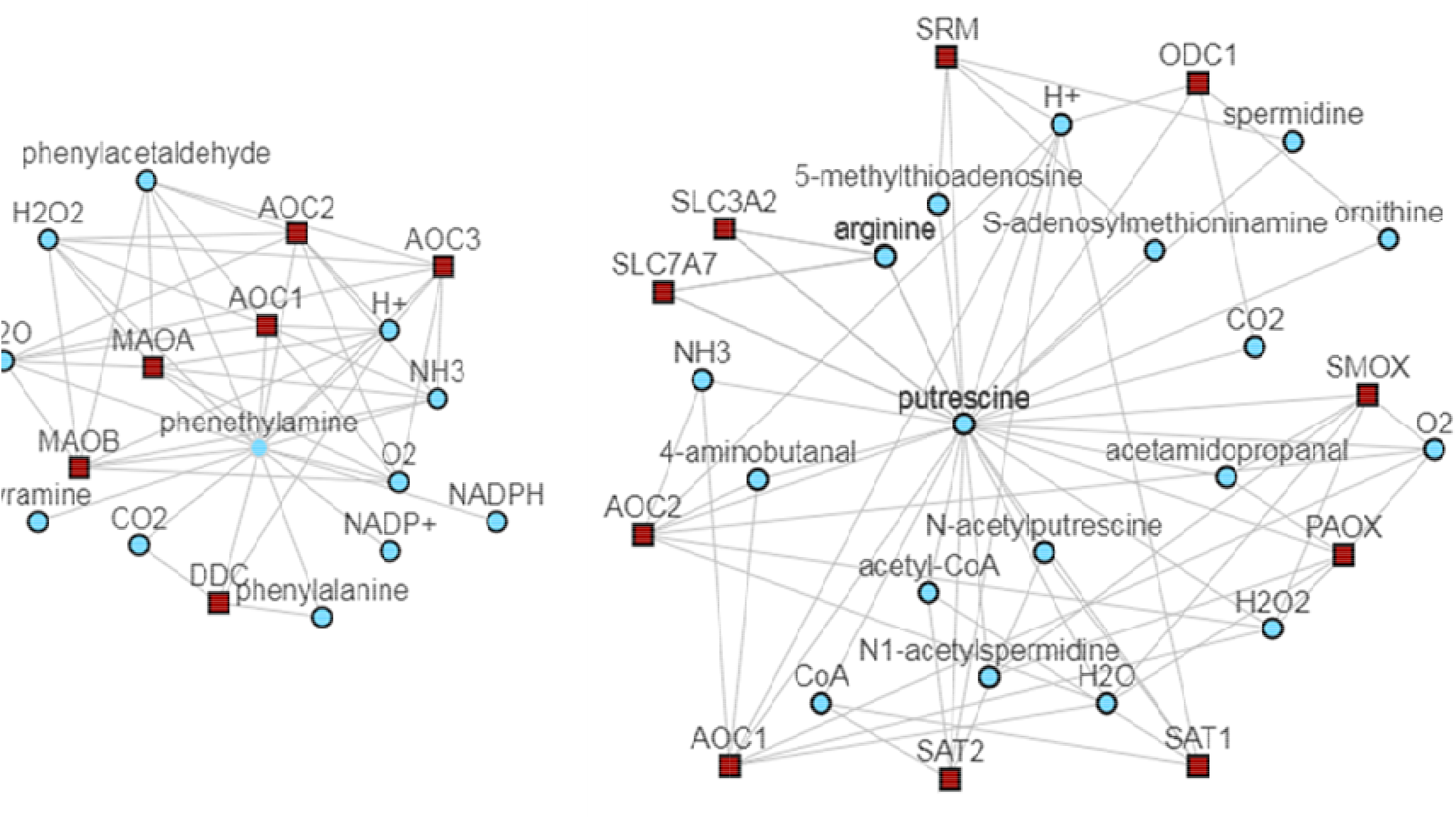
Metabolic Atlas analysis of interaction partners for PEA and putrescine. Many metabolites determined to be significantly altered in delirium-prone patients are closely linked to these two metabolites based on the metabolic network (Robinson et al. 2020).

### MAO oxidizes PEA and changes in MAO functional can explain observed differences in PEA in the two patient groups

Observed changes in PEA levels suggest changes in activity of MAO in delirium-prone patient cohort. MAOB is the only MAO protein found in platelets (for MAOA and MAOB protein expression please see Supplementary Fig. 3A). Other enzymes linked to PEA metabolism (Fig. 2) show significantly lower level of expression in tissues with particularly low expression in nervous system and thus cannot account for the change in PEA concentration in CSF (Supplementary Figure 3B)

### Analysis of published metabolomics data for COVID-19 patients shows significance difference in several metabolites that can be related to the changes in MAO function

In order to determine relationship between MAO and the related metabolites to the severity of SARS-CoV-2 response we have explored the dataset provided by (Shen et al. 2020) initially analysing the major metabolic differences between mild and severe COVID-19 cases. Patient information provided with the original publication does not include any data on possible delirium in these patients and we assumed, based on recent clinical studies (Mcloughlin et al. 2020) that patients with severe disease were more likely to have delirium than mild cases (with 60-70% in severe cases and 20-30% of hospitalized patients). For comparison we are providing major features selected using statistical, SAM methods (Fig. 3A) as well as machine learning – Random Forest methodology and SHAP analysis of feature (metabolite) contribution to the ML model (Fig. 3B).

**Fig. 3.**
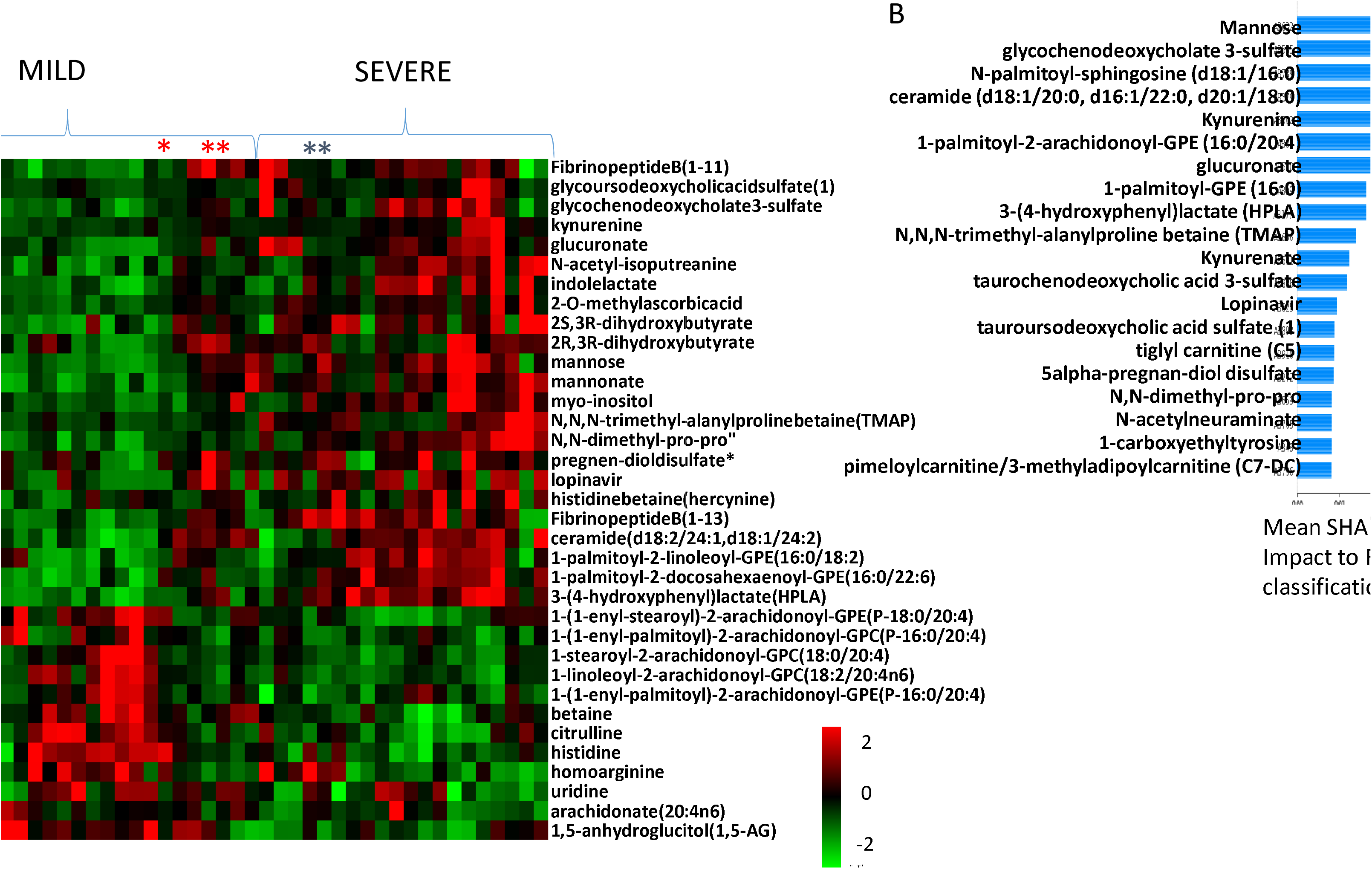
Analysis of major metabolic differences between Mild and Severe COVID-19 cases based on (Shen et al. 2020) dataset. A. SAM analysis of major features following sample and metabolite normalization; B. SHAP analysis of major contributors to Random Forest classification of mild *vs* severe cases.

The number of metabolites showing major concentration difference between severe and mild cases can be related to pathways involving MAO enzyme (see Supplementary Fig. 4 for all known direct interactions of MAOA and/or MAOB and metabolites). Specifically ratios of ceramide to sphingosine 1-phosphate (known as “sphingolipid rheostat”) is known to affect activity of MAO (Pchejetski et al. 2007). The ratio between these metabolites based on the COVID-19 patient data provided by (Shen et al. 2020) is shown in Fig. 4A and shows an increase with disease severity suggesting increasing activity of MAO. However, analysis of specific reaction partners that are related to MAO function shows a slight decrease of MAOB activity in severe cases (Fig. 4B) while MAOA activity (indicated with Fig. 4C reaction) is increasing in severe patient response.

**Fig. 4.**
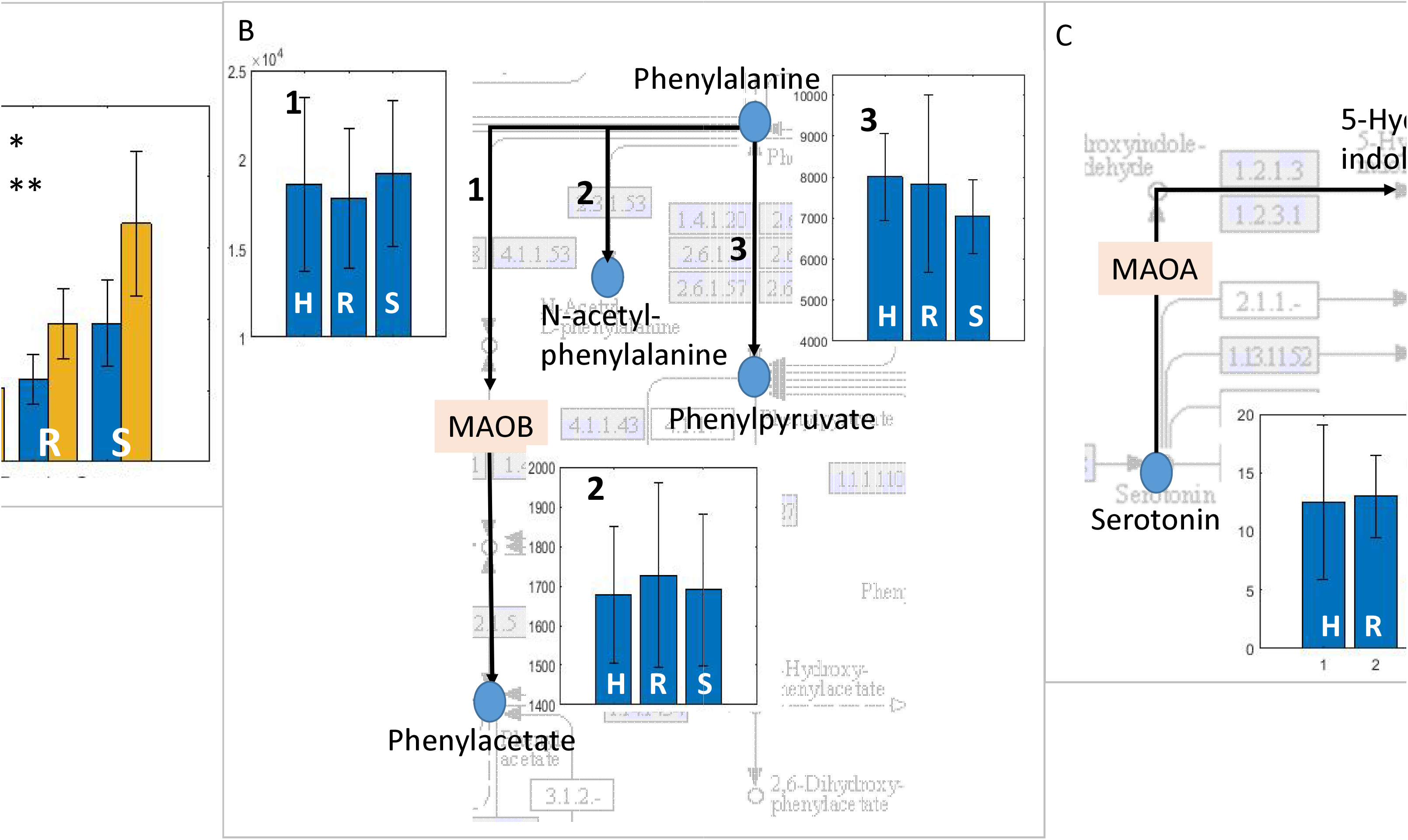
Median value and standard deviation of ratios between metabolites known to either activate MAO or to be part of metabolic activity of MAOA or B for healthy (H), mild (R) and severe (S) COVID-19 cases. A. Ratios of * ceramide (d18:1/20:0, d16:1/22:0, d20:1/18:0) / sphingosine 1-phosphate and ** ceramide (d18:2/24:1, d18:1/24:2) / sphingosine 1-phosphate. B. Part of Phenylalanine pathway with activity of MAOB showing ratio of 1. phenylalanine/phenylacetate; 2. phenylalanine/N-acetylphenylalanine and 3. phenylalanine/phenylpyruvate; C. part of tryptophan pathway where MAOA is used for serotonin metabolism. Shown is the median ratio of concentrations for serotonin/5-Hydroxyindolacetate.

### Major structural similarity between region of MAOB and ACE2 region binding to SARS-CoV-2 spike protein suggests possibility for MAOB-spike protein interaction

In order to investigate the possibility that the SARS-CoV-2 virus directly influences MAOB function the structures of MAOB and ACE2 (as a known binding target of SARS-CoV-2) were compared (Lan et al. 2020). Overall structural comparison between MAOB (PDB structure 1GOS) and ACE2 (PDB structure 6M0J), showed only 51% overall structural similarity. However the comparison of the ACE2 – spike protein binding region with MAOB resulted in 90% to 100% structure overlap. Further computational analysis of the spike-protein relation to MAOB protein is shown in Fig. 5.

**Fig. 5.**
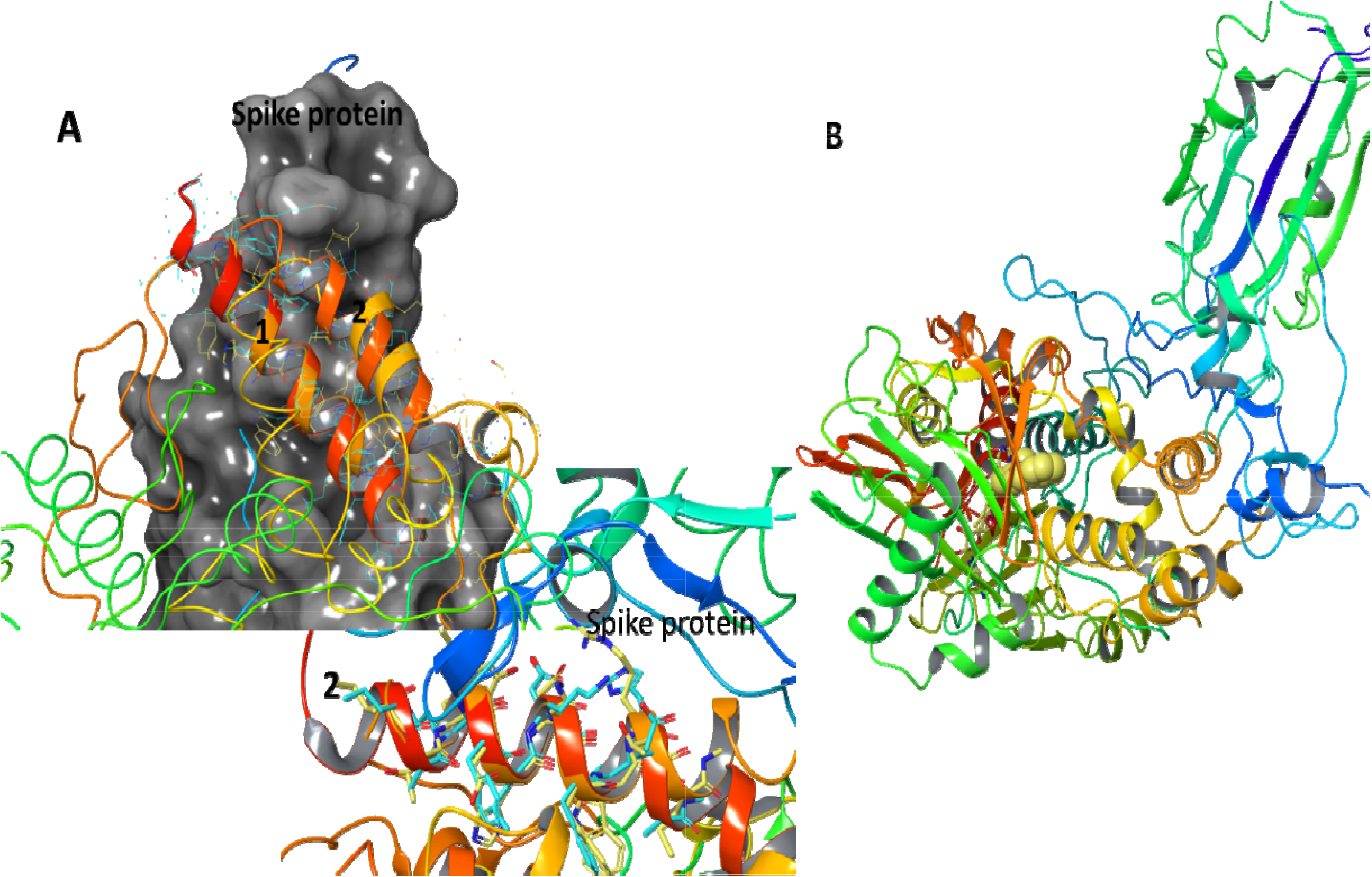
A. analysis of the overall structure similarity between ACE2 and MAOB using jFATCAT_flexible (Ye and Godzik, 2003) resulting in 6M0J Similarity: 42% 1GOS Similarity:51% P-value:6.67e-01. Further analysis of structure subsection structural alignment in the regions of ACE2 related to binding Spike protein are shown in 1 and 2 showing major similarity (Section 1 6M0J Similarity:100% 1GOS Similarity:100% Pvalue: 1.67e-02 Section 2 6M0J Similarity:74%; 1GOS Similarity:100% P-value:5.01e-01); B. Hypothetical binding between Spike protein and MAOB outlining the ligand location in the MAOB protein.

Specifically, analysis of the overall structure similarity between ACE2 and MAOB using jFATCAT_flexible (Ye and Godzik, 2003) resulted in ACE2 similarity of 42% and MAOB similarity of 51% with the P-value:6.67e-01. However, further analysis of subsection of ACE2 involved in binding to Spike protein indicated as 1 and 2 (Fig. 5A) shows major similarity (Section 1 6M0J Similarity:100% 1GOS Similarity:100% P-value:1.67e-02 Section 2 6M0J Similarity:74%; 1GOS Similarity:100% P-value:5.01e-01) suggesting possibility for interaction between MAOB and SARS-CoV-2 spike protein (Fig. 5A shows the association between MAOB and ACE2 overlapping regions and spike protein) leading to possibility for MAOB activity change in severe COVID-19 patients (Fig. 5B shows computational representation of the binding location of spike protein to MAOB indicating also the MAOB ligand).

### Computational evidence for Spike protein blocking entrance to the MAOB active site

Protein-protein docking analysis provides theoretical information about the most energetically beneficial binding orientation between proteins. In this analysis we have explored preferred arrangement with each chain of MAOB dimer acting as a receptor and SARS-CoV-2 spike protein viewed by the software as a ligand and *v.v*. Rigorous selection of the most energetically stable docking pose was selected amongst 70,000 different orientation of receptor and ligand proteins and calculated binding energy. The most stable docking form is provided in Fig. 6. The MAOB membrane bound residues have been excluded in the calculation of energy.

**Fig. 6.**
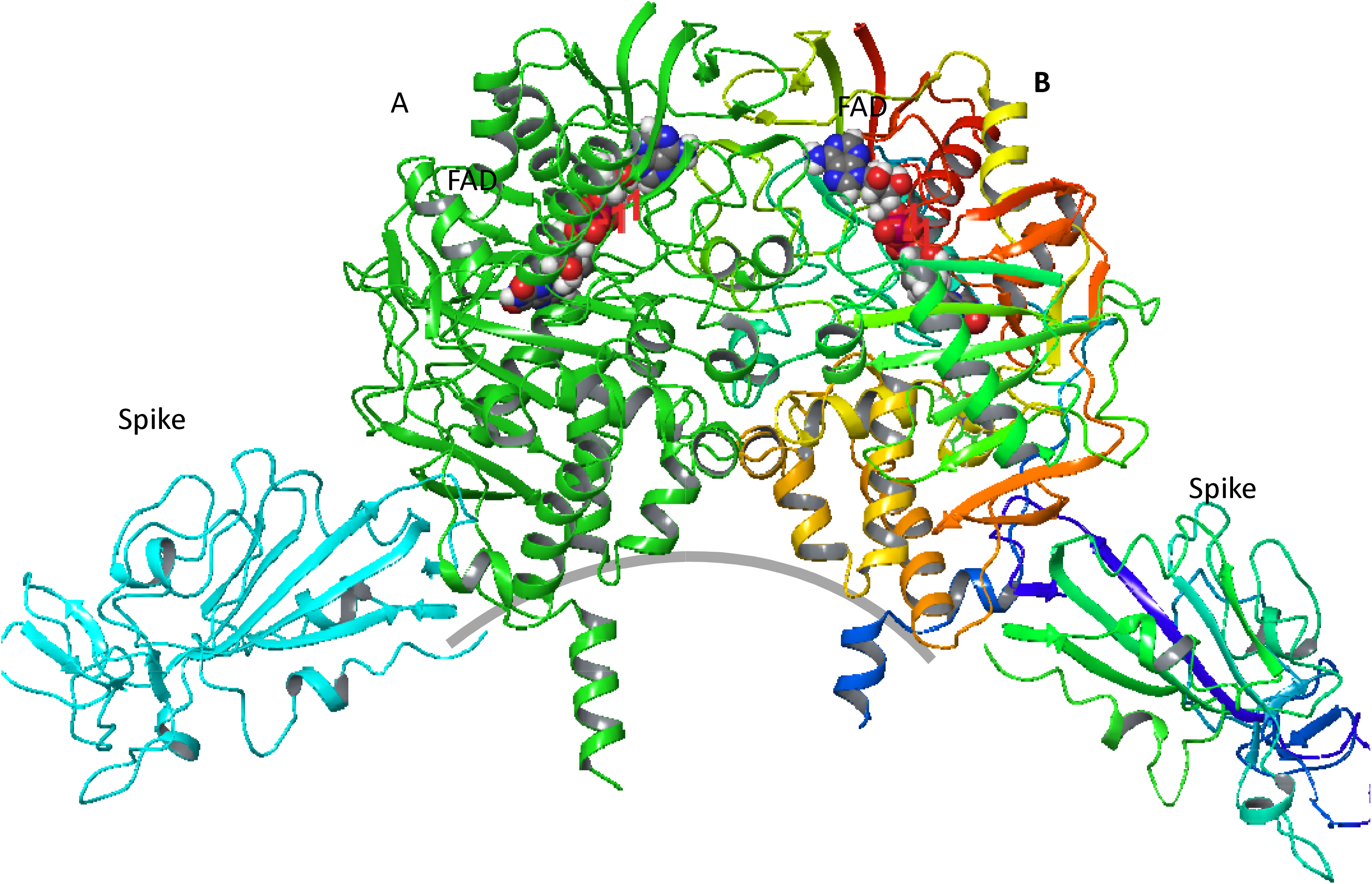
Computational Protein-Protein docking analysis of the energetically preferred binding location for Spike proteins to MAOB dimer (Scrodinger Inc.). Approximate location of the bilayer is indicated with a gray line. Binding energy calculation was performed for 70,000 consecutively selected positions of Spike protein as ligand and each chain of MAOB as a receptor. Shown are Spike proteins, MAOB chains as well as FAD – flavin adenine dinucleotide cofactor.

Potential energy of binding between spike proteins and two MAOB chains is calculated using OPLS3 force field -13424.67 kcal/mol (chain B) and -19073.44 kcal/mol (chain A). Resulting structure shows possible interference of spike protein with the recently presented membrane mediated substrate entrances to MAOB active site (Jones et al. 2019).

## Discussion

Delirium is an acute change in attention, awareness and cognition occurring as a result of a precipitants including medications, substance abuse, illness or surgery. Even routine surgical procedures, such as arthroplasty, are known to have post-operative delirium rates with 14% to 24% of elderly patients experiencing delirium after a routine surgery (Maldonado, 2013). A growing body of evidence links SARS-CoV-2 infection to delirium incidence (Alkeridy et al. 2020). This investigation generated and interrogated metabolomics data in order to identify the underlying metabolic perturbations associated with post-operative delirium and then explored possible SARS-CoV-2 related links to these pathways. A range of metabolite associations emerged (Fig. 1), but PEA was one of the most profoundly affected metabolites in patients who later experienced postoperative delirium. In delirium-prone individuals PEA concentrations are slightly reduced both in the CSF and blood plasma relative to the control group. Interestingly, for PEA an inverse relationship exists in these two compartments for these two groups with the significantly smaller PEA concentration in CSF relative to blood in delirium-prone cohort. Brain monoamines include common biogenic amines (dopamine, norepinephrine, and serotonin) and trace amines such as PEA (Xie et al. 2008). PEA has been shown to alter the serotonin transporter by interacting with trace amine-associated receptor 1 (TAAR1) (Xie et al. 2008). Activation of TAAR1 with PEA significantly inhibits uptake and induces efflux of dopamine and norepinephrine.

Observed alterations in PEA are highly plausible because monoamine oxidase (MAO), one of the key enzymes responsible for its metabolism, is a known target for treatment of a variety of neurological conditions including depression, Parkinson’s disease and recently Alzheimer’s disease (Yeung et al. 2019). MAO is an enzyme localised on the outer mitochondrial membrane and it preferentially degrades benzylamine and PEA. The MAO family of proteins oxidizes a number of different amine substrates including small-molecule monoamines, polyamines as well as modified amino acids in proteins, and directly influences number of different metabolites (Supplementary Fig. 4).

Two MAO subtypes exist: monoamine oxidase A (MAOA) which preferentially oxidizes biogenic amines such as 5-hydroxytryptamine (5-HT), norepinephrine and epinephrine, and monoamine oxidase B (MAOB) which performs oxidative deamination of biogenic and xenobiotic amines. MAOB is particularly important for the metabolism of neuroactive and vasoactive amines in the central nervous system and peripheral tissues. Expression of MAOB increases with age and is associated with increases in free radical damage and ROS formation. This in turn leads to a decrease in neuronal mitochondrial function and ultimately neurodegeneration (Kang et al. 2018) which is partly due to reduced PEA concentrations.

MAO inhibitors have been extensively developed and utilized for treatment of depression (Cleare et al. 2008, https://www.nice.org.uk/). A number of publications have also investigated the therapeutic effects of MAO inhibitors for other neurological conditions, such as Alzheimer’s disease, Parkinson’s disease, and depression (Yeung et al. 2019). Specifically, MAOB has been proposed as a possible therapeutic target for Alzheimer’s disease due to its association with aberrant GABA production, but it also has therapeutic relevance for Parkinson’s disease due to its role in dopamine depletion (Yeung et al. 2019, Park et al. 2019).

Utilization of MAO inhibitors (MAOI) for depression treatment has resulted in a number of side effects including: agitation, irritability, ataxia, movement disorders, insomnia, drowsiness, vivid dreams, cognitive impairment, and slowed speech, hallucinations and paranoid delusions (https://www.nice.org.uk/). Additionally, MAOI has been linked to an increasing suicide, pyrexia, delirium, hypoxia, hypertension and fatal intravascular coagulation (Fiedorowicz and Swartz, 2004; Tackley and Tregaskis, 1987; Thorp et al. 1997). MAOB is highly expressed in neurones, as well as platelets (Sandler and Reveley 1981) possibly explaining the observed effects of MAOB inhibitors on neurological state as well as blood based complications (Supplementary Fig. 3A shows protein expression of MAOA and MAOB in different human tissues). MAOB has been linked to the activity of platelets and dysfunction of nitric oxide synthase pathway observed in number of neurological diseases (recently reviewed in Leiter and Walker, 2020).

Previously reported links between HIV infection and changes in monoamine and acylcarnitine metabolites (as well as inflammatory markers) indicates that viral infection and inflammation can alter monoamine metabolism and mitochondrial energetics (Cassol et al. 2015). At the same time a number of side-effects previously listed for MAO inhibitors have also been observed in COVID-19 patients. One of the, as yet unresolved, effects of SARS-CoV-2 infection in a subgroup of patients is the development of a systemic coagulopathy and acquired thrombophilia characterized by a proclivity for venous, arterial and microvascular thrombosis (Becker, 2020). Additionally, severe cases of SARS-CoV-2 infection (where delirium is common) also experience low blood oxygen levels, elevated urea, acute renal dysfunction (Gulati et al. 2020) – all of which are symptomatic of MAOB inhibition overdose or drug side-effects such as anosmia, a known symptom of dopamine depletion in Parkinson’s disease. Differences in the activity of MAOB in surgical delirium-prone patient population was indicated by a change in the concentration of PEA as well as polyamines. Observed changes in the concentrations of carnitine derivatives in this patient cohort can be linked with previously established relationship between acetyl L-carnitine and dopamine through its effect on MAOB expression in the presence of anesthetic (Robinson et al. 2016). Significant concentration increases in severely affected COVID-19 patients is observed for both indolacetate and kynurenine as well as apparent increase in metabolising of serotonin in severe cases (Fig. 4) all part of tryptophan metabolism utilizing MAOA. Additionally, tryptophan plasma concentration in both mild and severe patients is significantly reduced relative to the healthy subjects (Supplementary Fig. 5) corroborated with the reduction in the concentration of arachidonate and related metabolites in severe relative to mild and both groups of infected patients relative to healthy subjects. Arachidonic acid is metabolized by activated platelets possibly leading to their aggregation. High risk groups for severe response to SARS-CoV-2 infection have known increased activity of MAO enzymes including: age, obesity, diabetes, heart condition (www.who.int). In addition to other symptoms, SARS-CoV-2 causes hematological changes which include reduced platelet count (Xu et al. 2020), platelet hyperactivity, changed gene expression in platelets (particularly in relation to protein ubiquitination), altered antigen presentation and also mitochondrial dysfunction (Manne et al. 2020). Furthermore, although neither ACE2 mRNA nor protein were detected in platelets, mRNA from the SARS-CoV-2 N1 gene was detected in the platelets of COVID-19 patients, suggesting that platelets may take-up SARS-COV-2 mRNA independently of ACE2 presence (Manne et al. 2020). Given the known role of MAO in a number of SARS-CoV-2 symptoms this prompted us to perform a computational analysis examining SARS-Co2 virus spike protein interactions with MAO. The preliminary computational structure comparison of MAOB and ACE2 protein performed here determined whether there is a possibility that the SARS-CoV-2 spike protein binds to MAOB (Fig. 5 and Supplementary Fig. 6-9). Although the sequence similarity between ACE2 and MAOB proteins is limited there is an almost coincident alignment and structural similarity in the region involved in SARS-CoV-2 spike protein binding (Supplementary Fig. 10). It is also important to point out that similar comparison between structures of ACE2 and AOC1, AOC2, AOC3 showed less than 30% overall similarity and no significant structural overlap with the ACE2 binding region for spike protein. Binding of the spike protein to MAOB could result in a change in either its enzymatic function, its post-translational modification or association with is protein partners including cell surface amino oxidases such as vascular adhesion protein 1 (VAP-1) (also known as AOC3) a known non-classical inflammation-inducible endothelial molecule (Salmi and Jalkanen, 2001).

The over-activity of MAOB can result in the observed PEA concentration decrease and the major changes observed in the polyamines as well as related amino acids in the delirium-prone patients. Pro-inflammatory stimuli, including cytokines lead to MAO-dependent increases in reactive oxygen species causing mitochondrial dysfunction (Deshwal et al. 2018). At the same time, interference with MAOB activity in subjects with overactive MAOB can lead to the side-effects observed in MAOB inhibition as well as SARS-CoV-2 infection. The metabolomic data and symptom similarities of delirium prone patients from a surgical setting and COVID-19 patients indicates potential dysfunction in MAOB. Dexamethasone recently shown to improve survival in severe COVID-19 cases (Horby et al., 2020) has been documented in the past to increase oxidative stress and the expression of MAOA (Ou et al., 2006) and MAOB (Tazik et al., 2009) in dopaminergic neurons (Johnson et al. 2010). Computational modeling defines a mechanism by which the spike protein directly binds to MAOB thereby interfering with its normal function and particularly affecting patients with increased MAOB expression. Detailed computational docking analysis shows strongest binding of spike protein to the region of MAOB recently proposed as the membrane mediated substrate entrance to the active site of MAOB (Jones et al. 2019). Further analysis is currently under way to explore in greater detail the role of MAOB in delirium and SARS-CoV-2 infection with further exploration of the effects of sex and other demographic, medical or drug utilization on the delirium related metabolic changes.

## Conclusion

Significant differences were observed in a number of mono- and polyamines which led us to investigate in some detail the relationship between the observed changes in operative delirium and delirium caused by SARS-CoV-2 and to propose a hypothetical relationship between monoamine oxidase and the SARS-CoV-2 spike protein. Experimental analysis of the relationship between delirium and SARS-CoV-2 and the possibility for spike-protein binding to monoamine oxidase is currently underway. Further research is required to establish what effect MAOB inhibitors might have on these pathways. There is no evidence at present to support the withholding of MAOB inhibitors in COVID-19 treatment.

## Data Availability

Data requests can be send to the authors.

## Acknowledgments

Many people contributed to the successful completion of this study. In particular we gratefully acknowledge the support of the anaesthetists, surgeons, theatre, ward and outcomes unit staff of Musgrave Park Hospital; Dr Seamus O’Brien who was key in the setup of the study; Mr John Conlon of the Centre for Experimental Medicine, QUB for technical laboratory support; Professor Chris Patterson of the Centre for Public Health for statistical support; Dr Rebecca Cairns and Ms Lauren Anderson for data inputting; Mrs Hazel Johnston and Mrs Eilish Armstrong for neuropsychology training; the Cheung family for support via the Siew Keok Chin scholarship. We especially acknowledge Dr Tim Mawhinney who was integral to the study set up and recruitment. Alzheimer’s Research UK (Metabolomic Analyses funded by Network Centre Pump Priming Grant). Cohort Study Funded by: Siew Keok Chin Scholarship (Philanthropic Funding). Belfast Arthroplasty Research Trust and Belfast Trust Charitable Funds.

MCC would like to thank Drs. Adrian Culf, Mary-Ellen Harper and Louis Borgeat for their reading of the manuscript and National Research Council Canada for supporting this research.

